# Association between sexual and reproductive health knowledge and protective sexual behaviors among young women in Nigeria

**DOI:** 10.1101/2022.02.16.22271074

**Authors:** David Aduragbemi Okunola, Oluwatobi Abel Alawode, Anthony Idowu Ajayi

## Abstract

**Background:** While the literature is replete with studies on young people’s risky sexual behaviors and their effects on HIV and STI acquisition, unintended pregnancy, and unsafe abortion, few studies have explored the practice of protective sexual behaviors and its association with sexual and reproductive health knowledge. We examined the association between sexual and reproductive health knowledge and protective sexual behaviors using data of young women in Nigeria.

**Methods:** Data of 8,995 young women (aged 15-24) who were neither married nor living with a partner were obtained from the 2018 Nigeria Demographic and Health Survey (NDHS) and analyzed in this study. Protective sexual behaviors were measured using questions exploring sexual fidelity, condom use, having multiple partners, and abstinence, while sexual and reproductive health knowledge was measured with questions on fertility, fecundity, contraceptives, and HIV prevention. We fitted multinomial logistic regression to examine the association between sexual and reproductive health knowledge and protective sexual behaviors.

**Results:** The median score on sexual and reproductive health knowledge was five. The majority of the sample (71.5%) had no sexual experience, 6.6% abstained from sex in the past year, 7.2% used condoms and practiced sexual fidelity, 1.0% used condoms but did not practice sexual fidelity, 12.7% did not practice sexual fidelity but used condoms while 0.8% neither used condom nor practiced sexual fidelity. After controlling for important covariates, sexual and reproductive health knowledge was positively associated with “condom use and sexual fidelity” (uRRR = 1.28; CI = 1.04-1.60) relative to “not using condoms nor practicing sexual fidelity.” However, the association between sexual and reproductive health knowledge and other categories of protective sexual behaviors, such as no sexual experience, abstinence in the past year, and sexual fidelity but no condom use, did not reach a significant level.

**Conclusion:** We found a high prevalence of protective sexual behavior among young women in Nigeria. Our study provides evidence in support of the association between sexual and reproductive health knowledge and a higher likelihood of practicing protective sexual behaviors. Intervention efforts can focus on providing sexual and reproductive health education to young people to equip them with information to safeguard their sexual health.

## Introduction

While the literature is replete with studies on young people’s risky sexual behaviors and their effects on unintended pregnancy, unsafe abortion, and HIV and STIs acquisition, few studies have explored their practice of protective sexual behaviors [1–8]. Protective sexual behavior is viewed as behaviors that reduce the risk of contracting sexually transmitted infections (STIs) and unintended pregnancies—including abstinence, sexual fidelity, and consistent use of condoms [9, 10]. Scholars tend to view young people’s sexual behaviors as dichotomous— as whether they use condoms or not—while neglecting the nuances inherent in their expression of their sexuality [10]. Such dichotomous views often erroneously characterize young people not using condoms but practicing sexual fidelity and using other protective methods (such as injectable, emergency contraceptives, and pre-exposure prophylaxis) as engaging in risky sexual behavior [2, 10]. The lack of focus on protective sexual behaviors often limits our understanding of the full spectrum of young people’s sexual behaviors. In addition, our limited understanding of factors driving young people to embrace protective sexual behaviors hinders our ability to develop effective programs to safeguard their sexual health.

Scholars who have studied protective sexual behaviors have shown that religiosity, HIV instrumental stigma beliefs, living with both parents, and adequate family support are positively associated with protective sexual behaviors. Ajayi and Okeke (2019) showed that young people from two-parent families who receive adequate family financial support are more likely to embrace protective sexual behaviors than their counterparts [10]. Somefun (2019), looking at the relationship between religiosity and sexual abstinence, observed that highly religious young people often display resilience in the face of risk and therefore tend to embrace abstinence [11]. Cort and Tu (2018) analyzed multi-country data from sub-Saharan Africa (SSA) and found that conservative HIV stigma attitudes are associated with lower risks of protective sexual behaviors [9]. However, there is limited evidence on the link between sexual and reproductive health knowledge and protective sexual behaviors. Although studies, mainly from the global north, have shown that teaching young people sexuality education, which encompasses building their reproductive health knowledge, is associated with improved sexual and reproductive health knowledge, attitudes, and behaviors [12–14]. Overall, there is limited evidence on how empowering young people with sexual and reproductive health knowledge could shape their sexual behaviors in SSA.

Reproductive empowerment through educating young people about sexual and reproductive health is transformative. It could expand individuals’ capacity to make informed decisions about their reproductive lives, amplify their ability to participate meaningfully in public and private discussions related to sexuality, reproductive health, and fertility, and act on their preferences to achieve desired reproductive outcomes, free from violence, retribution, or fear [15]. Although there is no consensus on how to measure reproductive empowerment, studies have suggested that women’s empowerment is associated with lower fertility, fertility desires, and higher contraceptive use among women [16–18]. These studies demonstrate the centrality of women’s empowerment for reproductive outcomes.

In this study, we propose that sexual and reproductive health knowledge is an important factor associated with young people’s practice of protective sexual behaviors, including sexual fidelity and condom use. Our assumption is premised on prior studies that have shown that teaching sexuality education is associated with positive sexual behaviors [12, 13]. SRHR knowledge is key to understanding different ways of safeguarding one’s sexual health. For example, knowledge of the ovulatory cycle and fecundability is important to prevent unintended pregnancy, and the knowledge of HIV transmission is key to preventing HIV [19, 20]. We examine our assumption using Nigeria’s 2018 demographic and health survey data of non-partnered young women aged 15 to 24 years. This age cohort has disproportionate exposure to adverse sexual and reproductive health outcomes, including new HIV infections and unintended pregnancies [21, 22]. Our study aligns with the global push for comprehensive sexuality education. Specifically, we compare the rate of protective sexual behavior among young people with a higher level of sexual and reproductive health knowledge to those with lower levels of knowledge. Our study contributes to the growing literature on factors influencing the practice of protective sexual behaviors among young people.

### Conceptual framework

We drew from International Center for Research on Women’s (ICRW) reproductive empowerment framework to understand through what mechanism sexual and reproductive health knowledge could impact protective sexual behaviors among young people [15]. The concept of empowerment is multidimensional, encompassing economic, social, cultural, legal, political, and psychological domains [23]. Pratley (2016) argues for the inclusion of a health-specific domain [24]. Similarly, the ICRW (2018) argues that reproduction is central to women’s empowerment and should be considered as a separate domain of empowerment [17]. Empowerment in the reproductive sphere entails both the agency and resources to make decisions and take action to attain sexual and reproductive health. Resources are enabling factors that act as catalysts for empowerment. Resources are not only material but also human and social resources, including comprehensive knowledge of SRHR, which serve to enhance the ability to exercise agency and choice. For example, knowledge of contraceptives, fecundity, ovulation, and HIV prevention methods can be quite important for young people’s sexual attitudes and behaviors.

For this reason, there is an emphasis on equipping young people with comprehensive sexuality education to enable them to make healthy relationship choices and express their agencies in reproductive decision-making [25, 26]. Several studies that aim to empower young people in the reproductive domains focus on designing curriculums to educate young people on reproductive health topics, including equitable gender norms, contraceptive methods and benefits, HIV/STI prevention methods, and healthy relationships, among others [27, 28]. Based on this conceptual framework, we argue that young people with reproductive health resources (comprehensive SRHR knowledge) would be more likely to express their agency and choice positively by embracing protective sexual behaviors. This leads us to hypothesize that young people’s level of reproductive empowerment is positively associated with the adoption and practice of protective sexual behaviors.

## Methods

### Data source

We used the individual recode file of the 2018 Nigeria Demographic and Health Survey (NDHS) publicly available through https://www.dhsprogram.com/data/available-datasets.cfm. The NDHS is a nationally representative periodic cross-sectional survey that provides information on demographic and health indicators. NDHS employed a stratified, two-stage cluster sampling technique to select eligible respondents (women aged 15-49), allowing for accurate and reliable estimates of demographic and health indicators that are representative at the national, regional, and state residential levels.

### Study population

This study considered only young women (aged 15-24) who were not partnered because this population is most at risk of contracting HIV and other STIs [21]. The NDHS interviewed 41,821 women (aged 15-49), out of which 8,995 (weighted sample) young women (aged 15-24) were neither married nor living with a partner were considered in this study. Limiting the dataset to only young women who were not partnered is consistent with a previous study on the subject [9].

### Measures

#### Outcome variable

The outcome variable in the study was protective sexual behavior. This variable is polytomous, with six different and mutually exclusive categories of sexual behaviors. Replicating the approach from a recent study on sub-Saharan Africa [9], information on young females related to (a) sexual experience, (a) sexual abstinence in the last 12 months, (c) condom use with the last and second-to last sexual partner, and (d) practice of sexual infidelity was combined to form the following six mutually exclusive categories of protective sexual behavior: (1) No sexual experience, (2) Abstinent over last 12 months, (3) Use condom and sexual fidelity, (4) Use condom and no sexual fidelity, (5) Sexual fidelity and no condom, and (6) No condom and no sexual fidelity.

#### Explanatory variable

The main explanatory variable of this study is sexual and reproductive health knowledge. We measured sexual and reproductive health knowledge as a variable using a composite score generated from dichotomous response options (yes (1) or no (0)) to questions about respondents’ knowledge of (a) ovulatory cycle, (a) postpartum fecundability, and (c) contraceptives. These questions were drawn from MacQuarrie’s (2021) multidimensional measure of youth empowerment, focusing on the reproductive health knowledge domain. Given that the questions in the domain are not exhaustive, we included the HIV prevention questions to have a more comprehensive measure of sexual and reproductive health knowledge. HIV prevention questions sought to know whether participants knew that: (a) one could reduce HIV risk by using condoms at every sex event and limiting to one uninfected partner with no other partner, (b) a healthy-looking person can have HIV, (c) mosquito bites cannot transmit HIV, and (d) one cannot become infected by sharing food with a person who has HIV. These responses (yes (1) or no (0)) were added to generate a composite score. Finally, the respondents’ composite scores on reproductive health knowledge and their comprehensive knowledge of HIV were summed to arrive at overall composite scores (ranging from 0 to 7) on reproductive health empowerment.

#### Covariates

We included individual and community level covariates in the study. At the individual level, we considered wealth status, which was created as terciles (poor, middle, and rich) from the original wealth quintile developed by the DHS program. Other individual covariates were age (15-19 years and 20-24 years), marital status (never married and divorced/separated), religion (Christianity and Islam/others), and ethnicity (Hausa, Igbo, Yoruba, and others). We also considered HIV stigma beliefs, which Cort and Tu (2018) have shown to be associated with protective sexual behaviors [9]. This was a composite score computed from the summation of responses (yes (1) or no (1)) to questions that examined: (a) whether female youths would buy vegetables from a vendor with HIV, (b) whether female youths would be ashamed if someone in their family had HIV, and (c) whether children with HIV should be allowed to attend schools with children without HIV. The resulting scores (ranging from 0 to 3) were trichotomized as (0) no stigma, (1) moderate stigma belief (1 for those who scored from 1 to 2), and high stigma belief (those who scored 3).

Community-level factors play important roles in youths’ sexual behaviors as such; we considered residence (urban and rural) and region of residence (Northcentral, North East, North West, South East, South-South, and South West) and the state-level HIV prevalence rates obtained from the 2019 Nigeria National HIV/AIDS Indicator and Impact Survey Technical report [29]. We grouped states with high HIV prevalence rates (2-5.9% prevalence) to form the high HIV prevalence cluster. States with HIV prevalence ranging from 1.1-1.9% were grouped into moderate HIV prevalence clusters, and those with less than 1% prevalence rate were grouped as low HIV prevalence clusters.

### Statistical analyses

All the variables were described using frequency, percentage distributions, and median. Multinomial logistic regression (MLR) was fitted at two stages to examine the relationship between the explanatory variable and protective sexual behavior. MLR was chosen for modeling protective sexual behavior because of its polytomous nature and nominal categories. In the first stage, the relationship between the explanatory variable and protective sexual behavior was separately examined using MLR: this produced unadjusted relative risk ratios. The final stage (the final model) contained all the covariates yielding adjusted relative risk ratios. The significance of the unadjusted relative risk ratios (uRRRs) and adjusted relative risk ratios (aRRRs) was tested against a 5% level of significance while their corresponding 95% confidence intervals were computed.

## Results

### Descriptive results

Table 1 presents the descriptive analysis of the variables in this study. The median year of education was 11 years. The majority of the young single women were less than 20 years old (72.51%) and never married (97.4%). More than half of the female youths were Christian (54.1%), in the wealthiest quintile (54.1%), urban residents (56.2%), and had low stigma belief (52.4%). Nearly one in five female youths (18.5%) resided in states with high HIV prevalence rates. The median score on reproductive health empowerment was five. The majority of the young single women (71.5%) had no sexual experience. About 7% of young women abstained from sex in the past year, 7.2% used condoms and practiced sexual fidelity, 1.1% used condoms but did not practice sexual fidelity, 12.7% did not use condoms but practiced sexual fidelity, and 0.8% of neither used condoms nor practiced sexual fidelity.

**Table 1.** Socio-demographic and behavioral characteristics of the respondents.

| Variables | Interval | % | Median | IQR | No of cases |
| --- | --- | --- | --- | --- | --- |
| **Outcome variable** |  |  |  |  |  |
| **Sexual Risk Behaviors** |  |  |  |  |  |
| No sexual experience |  | 71.47 |  |  | 6,429 |
| Abstinent over last 12months |  | 6.63 |  |  | 596 |
| Use condom & sexual fidelity |  | 7.24 |  |  | 652 |
| Use condom & no sexual fidelity |  | 1.07 |  |  | 96 |
| No condom & sexual fidelity |  | 12.75 |  |  | 1,147 |
| No condom & no sexual fidelity |  | 0.83 |  |  | 75 |
| **Explanatory variables** |  |  |  |  |  |
| **Reproductive health empowerment** | 0-7 |  | 5 | 3 |  |
| **Age** |  |  |  |  |  |
| 15-19 |  | 72.51 |  |  | 6,522 |
| 20-24 |  | 27.49 |  |  | 2,473 |
| **Marital status** |  |  |  |  |  |
| Never married |  | 97.42 |  |  | 8,763 |
| Divorced/separated |  | 2.58 |  |  | 232 |
| **Years of education** | 0-20 |  | 11 | 4 |  |
| **Wealth index** |  |  |  |  |  |
| Poor | 25.63 |  |  |  | 2,305 |
| Middle | 20.25 |  |  |  | 1,821 |
| Rich | 54.13 |  |  |  | 4,869 |
| **Religion** |  |  |  |  |  |
| Christianity |  | 54.13 |  |  | 4,869 |
| Islam / others |  | 45.87 |  |  | 4,126 |
| **Ethnicity** |  |  |  |  |  |
| Hausa/Fulani |  | 26.12 |  |  | 2,350 |
| Igbo |  | 18.07 |  |  | 1,626 |
| Yoruba |  | 18.24 |  |  | 1,641 |
| Others |  | 37.56 |  |  | 3,378 |
| **Residence** |  |  |  |  |  |
| Urban |  | 56.20 |  |  | 5,055 |
| Rural |  | 43.80 |  |  | 3,940 |
| **Region** |  |  |  |  |  |
| North Central |  | 14.98 |  |  | 1,347 |
| North East |  | 15.23 |  |  | 1,370 |
| North West |  | 22.31 |  |  | 2,007 |
| South East |  | 14.12 |  |  | 1,270 |
| South South |  | 13.78 |  |  | 1,240 |
| South West |  | 19.57 |  |  | 1,760 |
| **HIV belief stigma** |  |  |  |  |  |
| No stigma |  | 16.50 |  |  | 1,380 |
| Low stigma |  | 52.38 |  |  | 4,381 |
| High stigma |  | 31.12 |  |  | 2,603 |

| State-level HIV prevalence |  |  |  |  |  |
| --- | --- | --- | --- | --- | --- |
| Low |  | 43.02 |  |  | 3,870 |
| Moderate |  | 38.48 |  |  | 3,461 |
| High |  | 18.50 |  |  | 1,664 |

IQR = Interquartile range

#### Relationship between reproductive health empowerment and protective sexual behaviors

Table 2 presents the results from unadjusted (Model 1) and adjusted (Model 2) multinomial logistic regression models. In the first model, sexual and reproductive health knowledge was associated with protective sexual behavior. Specifically, an increase in sexual and reproductive health knowledge was associated with a 17% reduction in the probability of having no sexual experience (uRRR = 0.83; CI = 0.73-0.96) relative to not using condoms and not practicing sexual fidelity. However, after including covariates in Model 2, the strength of the relationship reduced, and the effect size was no longer significant. In contrast, the higher the sexual and reproductive health knowledge, the higher the probability of using condoms and practicing sexual fidelity (uRRR = 1.28; CI = 1.04-1.60) relative to not using condoms and not practicing sexual infidelity. The direction and strength of this relationship remained even with the inclusion of covariates in Model 2, thus providing evidence in support of our hypothesis that sexual and reproductive health knowledge is associated with protective sexual behavior. In Model 1, sexual and reproductive health knowledge was associated with a higher probability of using condoms and not practicing sexual fidelity (aRRR = 1.30; CI = 1.04-1.61) relative to not using condoms and not practicing sexual fidelity. However, in Model 2, the direction of the relationship remained while its strength reduced and was no longer significant. There was no significant relationship between sexual and reproductive health knowledge and abstinence in the last 12 months in both models, likewise not using condoms and practicing sexual fidelity.

**Table 2:** Multivariable multinomial logistic regression of protective sexual behavior on reproductive health empowerment.

| Protective sexual behaviors | Model 1<sup>a</sup><br>uRRR (CI) | Model 2<sup>b</sup><br>aRRR (CI) |
| --- | --- | --- |
| No sexual experience | 0.83(0.73,0.96)* | 0.96(0.79,1.16) |
| Abstinent over last 12 months | 1.00(0.87,1.15) | 1.03(0.85,1.26) |
| Use condom & sexual fidelity | 1.28(1.10,1.50)*** | 1.29(1.04,1.60)* |
| Use condom & no sexual fidelity | 1.30(1.04,1.61)* | 1.25(0.95,1.66) |
| No condom & sexual fidelity | 1.05(0.91,1.21) | 1.10(0.91,1.34) |

Base outcome =“no condom & no sexual fidelity”; \*\*\*P-value $\leq 0.001$ ; \*P < 0.05; CI: Confidence Interval; RC = Reference Category; <sup>a</sup>No covariate present; <sup>b</sup>covariates present: age, marital status, education level, wealth status, religion, ethnicity, residence, region, HIV stigma beliefs, and state-level HIV prevalence; uRRR: unadjusted relative risk ratio; aRRR: adjusted relative risk ratio

## Discussion

Our study examined the patterns of protective sexual among young women (aged 15 to 24) and the role of sexual and reproductive health knowledge. We found that most young women in the study setting engaged in protective sexual behaviors, including abstinence, sexual fidelity, and condom use. However, nearly one in 100 young women did not use condoms and did not practice sexual fidelity. While this rate may appear negligible, it is, in fact, a substantial number of young people in Nigeria practicing no condoms and no sexual fidelity, considering the high young women population in Nigeria [30]. Overall, our findings suggest that the practice of protective sexual behavior is high among young women in Nigeria, which is consistent with past studies [10, 11].

In support of our hypothesis, we found a significant positive association between sexual and reproductive health knowledge and condom use and sexual fidelity. However, the effect size of the association between sexual and reproductive health knowledge and condom use but no sexual fidelity no longer reached a significant threshold after controlling for important covariates, even though the direction of the effect remains. Overall, our study presents robust evidence of the importance of empowering young people with sexual and reproductive health information. When young people are empowered with information on sexual and reproductive health, they are more likely to embrace protective sexual behaviors and make healthy choices about their reproductive health [25, 31]. Even though there is limited evidence on the association between sexual and reproductive knowledge and protective sexual behavior, our finding is consistent with previous studies reporting a positive association between measures of empowerment or agency and contraceptive use [32–34]. Our finding is consistent with studies that have reported the association between sexuality education and young people’s sexual behavior [14, 35]. While young people receive limited sexuality education in Nigerian schools, information on sexuality is available online and through the media. Many young women learn about sexual and reproductive health, including contraceptive methods, especially after becoming sexually active [36, 37]. However, those who lack access to sexual and reproductive health information or those exposed to wrong information are at risk of not adequately protecting themselves against unintended pregnancies and STIs. Our study underscores the need for targeted sexuality education interventions for young women to safeguard their sexual health.

### Implications of the study

Our study findings can inform programs and interventions targeting young people with sexual and reproductive health information. Educating young people about fecundability, contraceptive use, HIV prevention methods, and other sexual and reproductive health topics is critical. Investing in young people’s sexual and reproductive health skills and resources is crucial for increasing their reproductive health empowerment (agency). Such investment will yield benefits by increasing their ability to practice protective sexual behaviors that could help them make healthy life choices, including sexual fidelity and contraceptive use, and prevent HIV, STIs, and unintended pregnancies.

### Study strengths and limitations

This study has made important contributions to the literature on protective sexual behaviors. The data used is nationally representative, allowing for robust analysis and generalization. However, it is important to highlight a few limitations of the study. Due to the limitation of secondary data used in this study, our sexual and reproductive health knowledge measure is neither exhaustive nor comprehensive. The study’s cross-sectional nature means we could not draw a causal link between sexual and reproductive health knowledge and protective sexual behaviors. Also, self-reporting of sensitive issues such as sex, sexual fidelity, and contraceptive use is fraught with social desirability bias, suggesting our study could have overestimated protective sexual behaviors prevalence.

Nonetheless, our study makes important contributions to the emerging literature on the topic in Nigeria. It is one of the first studies to focus on sexual and reproductive health knowledge and protective sexual behaviors. Future studies should build on our study by developing a multidimensional measure for sexual and reproductive health knowledge and examine how it influences protective sexual behaviors.

### Conclusion

We found a high prevalence of protective sexual behavior among young women in Nigeria. Our study provides evidence on the association between sexual and reproductive health knowledge and the likelihood of practicing protective sexual behaviors. Based on our findings, we conclude that a higher level of sexual and reproductive health knowledge can increase the likelihood of adopting sexual protective sexual practices such as sexual fidelity and the use of condoms. Intervention efforts can focus on providing sexual and reproductive health education to young people to equip them with information to safeguard their sexual health.

## Data Availability

The data underlying the results presented in the study are available from https://www.dhsprogram.com/data/available-datasets.cfm

## Acknowledgement

We thank measure DHS for making the data analyzed in this study publicly available and David Cort for supporting our data analysis.

## Ethical consideration

The Nigeria Ministry of Health’s ethics committee granted ethics approval for the 2018 NDHS. Since we analyzed de-identified dataset that is publicly available, we do not require another ethics authorization.

## Authors’ contributions

DAO, OAA and AIA conceptualized this study. DAO conducted the analysis with inputs from AIA. All authors contributed to drafting and reviewing the manuscript.

## Data sharing

Data analyzed in this study is available via https://www.dhsprogram.com/data/available-datasets.cfm

## References

1. Adeboye A, Yongsong Q, James N. Risky Sexual Behavior and Knowledge of HIV/AIDS among High School Students in Eastern Cape South Africa. J Hum Ecol. 2016;53(3):194–204.

2. Ali M, Ajilore O. Risky sexual behavior and African-American youth: What is the role of family structure? J Health Behav & Pub Health 2011;1(1):30–40

3. Imaledo JA, Peter-Kio OB, Asuquo EO. Pattern of risky sexual behavior and associated factors among undergraduate students of the University of Port Harcourt, Rivers State, Nigeria. Pan Afr Med J. 2012;12(1).

4. Kebede A, Molla B, Gerensea H. Assessment of risky sexual behavior and practice among Aksum University students, Shire Campus, Shire Town, Tigray, Ethiopia, 2017. BMC Res Notes. 2018;11(1):88.

5. Oluwatoyin F, Oyetunde M. Risky sexual behavior among secondary school adolescents in Ibadan North Local Government Area, Nigeria. JNHS. 2014;3:34–44.

6. Othieno CJ, Okoth R, Peltzer K, Pengpid S, Malla LO. Risky HIV sexual behaviour and depression among University of Nairobi students. Ann Gen Psychiatry. 2015;14(1):16.

7. Ritchwood TD, Ford H, DeCoster J, Sutton M, Lochman JE. Risky sexual behavior and substance use among adolescents: A meta-analysis. Child Youth Serv Rev. 2015;52:74–88.

8. Tsutsumi A, Izutsu T, Matsumoto T. Risky sexual behaviors, mental health, and history of childhood abuse among adolescents. Asian J Psychiatr. 2012;5(1):48–52.

9. Cort DA, Tu HFJSS, Medicine. Safety in stigmatizing? Instrumental stigma beliefs and protective sexual behavior in Sub-Saharan Africa. Soc Sci Med. 2018;197:144–52.

10. Ajayi AI, Okeke SR. Protective sexual behaviours among young adults in Nigeria: influence of family support and living with both parents. BMC Public Health. 2019;19(1):983. doi: 10.1186/s12889-019-7310-3.

11. Somefun OD. Religiosity and sexual abstinence among Nigerian youths: does parent religion matter? BMC Public Health. 2019;19(1):416.

12. Kirby DB. The impact of abstinence and comprehensive sex and STD/HIV education programs on adolescent sexual behavior. Sex Res Social Policy. 2008;5(3):18–27.

13. Fonner VA, Armstrong KS, Kennedy CE, O’Reilly KR, Sweat MD. School based sex education and HIV prevention in low-and middle-income countries: a systematic review and meta-analysis. PLOS ONE. 2014;9(3):e89692.

14. Cheedalla A, Moreau C, Burke AE. Sex education and contraceptive use of adolescent and young adult females in the United States: an analysis of the National Survey of Family Growth 2011–2017. Contraception: X. 2020;2:100048.

15. Edmeades J, Hinson L, Sebany M, Murithi L. A conceptual framework for reproductive empowerment: Empowering individuals and couples to improve their health (Brief). Washington, DC: International Center for Research on Women. 2018.

16. James-Hawkins L, Peters C, VanderEnde K, Bardin L, Yount KM. Women’s agency and its relationship to current contraceptive use in lower-and middle-income countries: A systematic review of the literature. Glob Public Health. 2018;13(7):843–58.

17. Upadhyay UD, Dworkin SL, Weitz TA, Foster DG. Development and validation of a reproductive autonomy scale. Stud Fam Plann. 2014;45(1):19–41.

18. Upadhyay UD, Gipson JD, Withers M, Lewis S, Ciaraldi EJ, Fraser A, et al. Women’s empowerment and fertility: a review of the literature. Soc Sci Med. 2014;115:111–20.

19. Long L, Yuan T, Wang M, Xu C, Yin J, Xiong C, et al. Factors associated with condom use among male college students in Wuhan, China. PLOS ONE. 2012;7(12):e51782.

20. Frost JJ, Lindberg LD, Finer LB. Young adults contraceptive knowledge, norms and attitudes: associations with risk of unintended pregnancy. Perspect Sex Reprod Health. 2012;44(2):107–16.

21. Organization WH. Global health sector strategy on HIV 2016-2021. Towards ending AIDS. World Health Organization, 2016.

22. Adeniyi OV, Ajayi AI, Moyaki MG, Ter Goon D, Avramovic G, Lambert J. High rate of unplanned pregnancy in the context of integrated family planning and HIV care services in South Africa. BMC Health Serv Res. 2018;18(1):140.

23. Malhotra A, Schuler SR. Women’s empowerment as a variable in international development. Measuring empowerment: Cross-disciplinary perspectives. 2005;1(1):71–88.

24. Pratley P. Associations between quantitative measures of women’s empowerment and access to care and health status for mothers and their children: a systematic review of evidence from the developing world. Soc Sci Med. 2016;169:119–31.

25. Haberland N, Rogow D. Sexuality education: emerging trends in evidence and practice. J Adolesc Health. 2015;56(1):S15–S21.

26. Goldman JD. A critical analysis of UNESCO’s International Technical Guidance on school-based education for puberty and sexuality. Sex Educ. 2012;12(2):199–218.

27. Austrian K, Soler-Hampejsek E, Kangwana B, Maddox N, Wado YD, Abuya B, et al. Adolescent Girls Initiative–Kenya: Endline evaluation report. 2020.

28. Austrian K, Soler-Hampejsek E, Kangwana B, Wado Y, Abuya B, Maluccio J. Impacts of two-year multi-sectoral interventions on young adolescent girls’ education, health and economic outcomes: Adolescent Girls Initiative-Kenya randomized trial. 2020.

29. Federal Ministry of Health N. Nigeria HIV/AIDS Indicator and Impact Survey report (NAIIS) 2018: Technical Report Abuja, Nigeria: Federal Ministry of Health, Nigeria., 2019.

30. United Nations, Department of Economic and Social Affairs, Population Division. 2019 Revision of World Population Prospects. New York: United Nations, 2019 0897149726.

31. UNESCO. The Journey towards Comprehensive Sexuality Education. Paris, France: UNESCO, 2021.

32. Do M, Kurimoto N. Women’s empowerment and choice of contraceptive methods in selected African countries. Int Perspect Sex Reprod Health. 2012:23–33.

33. Vizheh M, Muhidin S, Behboodi Moghadam Z, Zareiyan A. Women empowerment in reproductive health: a systematic review of measurement properties. BMC Womens Health. 2021;21(1):1–13.

34. Viswan SP, Ravindran TS, Kandala N-B, Petzold MG, Fonn S. Sexual autonomy and contraceptive use among women in Nigeria: findings from the demographic and health survey data. Int J Womens Health. 2017;9:581.

35. Boti N, Hussen S, Shegaze M, Shibru S, Shibiru T, Zerihun E, et al. Effects of comprehensive sexuality education on the comprehensive knowledge and attitude to condom use among first-year students in Arba Minch University: a quasi-experimental study. BMC Res Notes. 2019;12(1):1–7.

36. Ajayi AI, Nwokocha EE, Adeniyi OV, Ter Goon D, Akpan W. Unplanned pregnancy-risks and use of emergency contraception: a survey of two Nigerian Universities. BMC Health Serv Res. 2017;17(1):382. doi: 10.1186/s12913-017-2328-7.

37. Ajayi AI, Olamijuwon EO. What predicts self-efficacy? Understanding the role of sociodemographic, behavioural and parental factors on condom use self-efficacy among university students in Nigeria. PLOS ONE. 2019;14(8):e0221804.

